# Myocardial characteristics as the prognosis for COVID-19 patients

**DOI:** 10.1101/2020.05.06.20068882

**Authors:** Jianguo Zhang, Daoyin Ding, Can Cao, Jinhui Zhang, Xing Huang, Peiwen Fu, Guoxin Liang, Wenrong Xu, Zhimin Tao

## Abstract

**Background:** Amid the crisis of coronavirus disease 2019 (COVID-19) caused by the severe acute respiratory syndrome coronavirus 2 (SARS-CoV-2), front-line clinicians in collaboration with backstage medical researchers analyzed clinical characteristics of COVID-19 patients and reported the prognosis using myocardial data records upon hospitalization.

**Methods:** We reported 135 cases of laboratory-confirmed COVID-19 patients admitted in The First People’s Hospital of Jiangxia District in Wuhan, China. Demographic data, medical history, and laboratory parameters were taken from inpatient records and compared between patients at the Intensive Care Unit (ICU) and non-ICU isolation wards for prognosis on disease severity. In particular, survivors and non-survivors upon ICU admission were compared for prognosis on disease mortality.

**Results:** For COVID-19 patients, blood test results showed more significantly deranged values in the ICU group than those in non-ICU. Among those parameters for ICU patients, myocardial variables including troponin T, creatine kinase isoenzymes, myoglobin, were found significantly higher in non-survivors than in survivors.

**Conclusions:** Upon hospitalization abnormal myocardial metabolism in COVID-19 patients could be prognostic indicators of a worsened outcome for disease severity and mortality.

## Introduction

On December 2019, a novel coronavirus emerged and provoked a global spreading of pneumonia diseases [1-4]. The disease was later named as coronavirus disease 2019 (COVID-19) [5]. The pathogen responsible for the COVID-19 was discovered as the severe acute respiratory syndrome coronavirus 2 (SARS-CoV-2). As of April 18, 2020, over 2 million cases of COVID-19 infection were confirmed globally, constituting an unprecedented public health emergency [6]. With no effective treatment available, the mortality rate of COVID-19 has been estimated ~ 5.4% [6].

Upon inpatient admission, poor prognosis was associated with elderly age, high Sequential Organ Failure Assessment (SOFA) score, and augmented D-dimer level of COVID-19 patients [7]. Since the epidemic strike of COVID-19, respiratory and pulmonary systems have been heavily investigated to enhance understanding towards this devastating infectious disease, leaving cardiovascular parameters of patients a less explored area. Here we reported 135 laboratory-confirmed COVID-19 patients in Wuhan, China, by analyzing clinical data upon their hospital admission with a linkage to their final outcomes. Through this study by comparing clinical characteristics among non-severe, severe and deceased groups of patients, we aimed to reveal the relationship between myocardial characteristics and disease prognosis of COVID-19.

## METHODS

### Study Design

The study was approved by The First People’s Hospital of Jiangxia District (TFPHJD) (Approval No. 2020021) in Wuhan, China. With outbreak of unknown viral pneumonia cases in Wuhan, the dispatch of medical team from The Affiliated Hospital of Jiangsu University (TAHJU) was sent to execute medical reinforcement in TFPHJD on Jan 27, 2019. A total of 135 COVID-19 patients were hospitalized in TFPHJD between February 1 and March 15, 2020, and data were collected including 30 patients in the Intensive Care Unit (ICU) and 105 patients in non-ICU isolation ward. Written consent was waived by Ethics Commission of TFPHJD due to the emergency of a major infectious disease.

### Procedure

All COVID-19 patients were received at TFPHJD and diagnosed by following a standard procedure [8]. The confirmed patients were treated with antiviral drugs, including oseltamivir, arbidol, and ribavirin. For the severe patients who were admitted into ICU by following the published criteria [8], they typically developed hypoxemia, dyspnea, and even respiratory failure requiring respiratory support or invasive mechanical ventilation, and usually had complications such as liver and kidney injuries, which required blood purification. They were receiving antibiotic treatment (sulperazone, linezolid), antifungal therapy (fluconazole, caspofungin), corticosteroid therapy, respiration-assisted ventilation, continuous renal replacement therapy. For all patients, blood cell analysis was detected by automated hematology analyzer (SYSMEX 800i, Japan), and the biochemical indicator was analyzed (Toshiba TAB2000, Japan). The biochemical markers of myocardial injury were measured (Roche Cobas 6000 Analyzer, Switzerland).

### Date Collection

Demographic data, medical history and clinical characteristics including symptoms, chest computed tomography (CT) scans, myocardial biomarkers, hematological and biochemical tests, were obtained at TFPHJD. The data access and the study were approved by Ethics Commission of TFPHJD.

### Statistical Analysis

The categorical variables were described as frequency rates and percentages, and continuous variables were applied to describe the median and quartile range (IQR) values. Comparison of continuous variables between two groups was analyzed with unpaired Student’s t test or Mann-Whitney test, as appropriate. Repeated measurements (non-normal distribution) were used following a generalized linear mixed model. χ^2^ test was used to compare the proportion of categorical variables, and the Fisher exact test was employed when data was limited. All statistical analyses were performed using SPSS (statistical package for social sciences) version 13.0 software (SPSS Inc.). A two-sided α of less than 0.05 was considered statistically significant unless otherwise specified.

## RESULTS

### Baseline Characteristics of 135 COVID-19 Patients

Admitted at TFPHJD, a total of 135 COVID-19 patients aged 17 to 89 years old were included in this study. The median age was 56.0 years (IQR 42.0-68.0), and 67 (49.6%) of them were male (Table 1). 30 (22.2%) patients were later transferred into the ICU based on the severity of disease development [8]. The other 105 (77.8%) remained in the non-ICU isolation wards until recovery and discharge from the hospital. Non-ICU patients were between 17 to 89 years old with a median age of 54.0 years (IQR 41.0-67.5), where 52 (49.5%) of them were male. In comparison, ICU patients were between 36 to 88 years old with a median age of 64.0 years (IQR 50.0-72.3), where 15 (50.0%) of them were male. This result indicated that the median age of patients in ICU was significantly higher than that in non-ICU ward (P<0.05), while gender difference had little correlation to the transmission and development of the COVID-19 diseases.

**Table 1.** Demographic data, medical history and clinical symptoms of 135 COVID-19 patients.

|  | **Total (n=135)** | **ICU (n=30)** | **Non-ICU (n=105)** | **P value** |
| --- | --- | --- | --- | --- |
| Age, year | 56.0 (42.0-68.0) | 64.0 (50.0-72.3) | 54.0 (41.0-67.5) | 0.013 |
| Gender, male | 67 (49.6%) | 15 (50.0%) | 52 (49.5%) | 0.963 |
| **Comorbidities** | **46 (34.1%)** | **17 (56.7%)** | **29 (27.6%)** | **0.003** |
| Hypertension | 28 (20.7%) | 11 (36.7%) | 17 (16.2%) | 0.015 |
| Diabetes | 16 (11.9%) | 4 (13.3%) | 12 (11.4%) | 0.754 |
| Cardiovascular diseases | 4 (3.0%) | 1 (3.3%) | 3 (2.9%) | 1.000 |
| Bronchitis | 3 (2.2%) | 3 (10.0%) | 0 (0.0%) | 0.010 |
| Hypothyroidism | 2 (1.5%) | 0 (0.0%) | 2 (1.9%) | 1.000 |
| Hepatitis B | 2 (1.5%) | 2 (6.7%) | 0 (0.0%) | 0.048 |
| Renal dysfunction/failure | 2 (1.5%) | 2 (6.7%) | 0 (0.0%) | 0.048 |
| Renal calculi | 1 (0.7%) | 1 (3.3%) | 0 (0.0%) | 0.222 |
| Acute pancreatitis | 1 (0.7%) | 0 (0.0%) | 1 (1.0%) | 1.000 |
| Cholecystitis | 1 (0.7%) | 0 (0.0%) | 1 (1.0%) | 1.000 |
| Gallstone | 1 (0.7%) | 0 (0.0%) | 1 (1.0%) | 1.000 |
| Gout | 1 (0.7%) | 0 (0.0%) | 1 (1.0%) | 1.000 |
| Epilepsy | 1 (0.7%) | 1 (3.3%) | 0 (0.0%) | 0.222 |
| Intracerebral hemorrhage | 1 (0.7%) | 1 (3.3%) | 0 (0.0%) | 0.222 |
| **Clinical symptoms** |  |  |  |  |
| Cough | 135 (100.0%) | 30 (100.0%) | 105 (100.0%) | N/A |
| Fever | 128 (94.8%) | 30 (100.0%) | 98 (93.3%) | 0.348 |
| Fatigue | 82 (60.7%) | 20 (66.7%) | 62 (59.0%) | 0.451 |
| Chest pain | 46 (34.1%) | 14 (46.7%) | 32 (30.4%) | 0.099 |
| Diarrhea | 32 (23.7%) | 8 (26.7%) | 24 (22.9%) | 0.665 |
| Vomiting | 24 (17.8%) | 6 (20.0%) | 18 (17.1%) | 0.718 |
| Dyspnea | 10 (7.4%) | 6 (20.0%) | 4 (3.8%) | 0.008 |

Among 135 hospitalized patients, 89 (65.9%) had no known acute or chronic diseases, showing their healthy history prior to SARS-CoV-2 infection. For patients with comorbidity, 28 (20.7%) had hypertension, 16 (11.9%) diabetes, and 4 (3.0%) cardiovascular diseases. Less common comorbidity could be found as shown in Table 1. Among those co-existing medical conditions, shown as [ICU versus (vs.) non-ICU patients], hypertension [11 (36.7%) vs.17 (16.2%)] put patients at the highest risk to infection and once infected those patients could develop worsened conditions. 56.7% of ICU patients had one comorbidity or more, whereas only 27.6% non-ICU patients had at least one comorbidity. We calculated the individual comorbidity percentage in ICU and non-ICU patients, and plotted them versus each other as shown in Figure 1A. Dotted diagonal (in red) indicated a hypothetically equal percentage between the two groups. As a result, hypertension stayed the farthest away from the diagonal, showing this comorbidity possibly the highest risk factor to cause COVID-19 patients on further critical condition. Following the same analysis, the other co-existing diseases, such as diabetes, bronchitis, and renal dysfunction/failure, make up probable risk factors to have negative impact on COVID-10 patients, which may aggravate the disease severity after virus infection.

**Figure 1.**
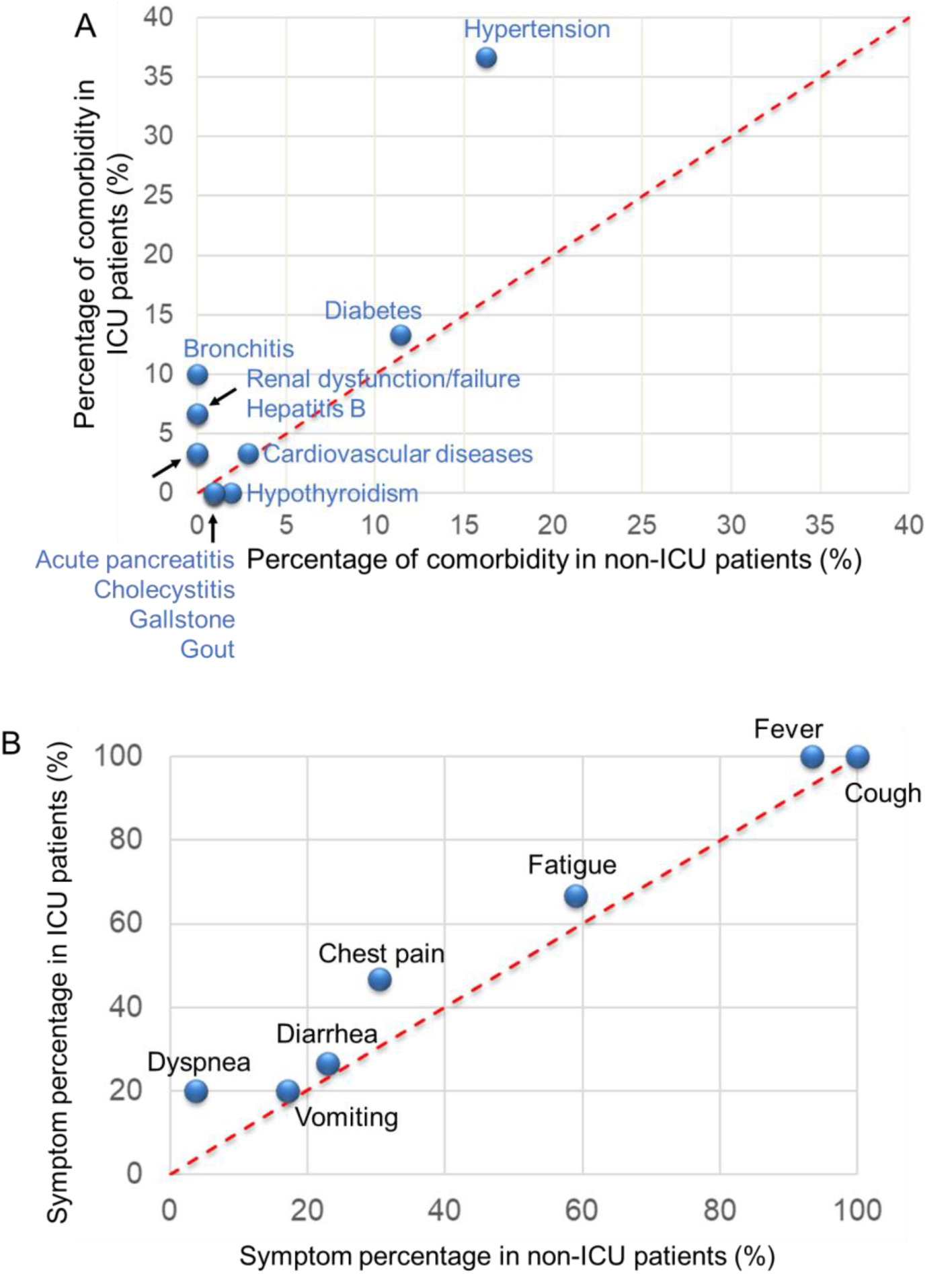
Comorbidity percentage (A) or symptom percentage (B) in ICU and non-ICU patients was plotted versus each other. Diagonal (red, dotted) indicated a hypothetically equal percentage between the two groups. Overlapping coordinates were pointed by an arrow with following descriptions.

At the onset of COVID-19 illness, the most common symptoms appeared in 135 patients were cough (135 patients, 100%), fever (128, 94.8%), fatigue (82, 60.7%), chest pain or distress (46, 34.1%), diarrhea (32, 23.7%), vomiting (24, 17.8%), and dyspnea (10, 7.4%) (Table 1). Notably, 5.2% later-confirmed positive patients experienced no fever. For patients showing dyspnea, 6 were transferred to ICU while the other 4 stayed in non-ICU isolation until fully recovered. Each symptom had a higher incidence in ICU than in non-ICU. We calculated the percentage of the above symptoms in ICU and non-ICU groups, respectively, and plotted the symptom percentage in ICU vs. that in non-ICU. As shown in Figure 1B, the diagonally dotted line indicated a hypothetically equal symptom percentage between two groups. This figure depicted that the later-admitted ICU patients were more likely to show initial symptoms compared with non-ICU patients, as most points that belong to ICU patients scattered above the diagonal.

### Blood Abnormality of COVID-19 Patients Upon Hospitalization

We performed the laboratory tests of COVID-19 patients on their first day of hospitalization, and typical parameters indicating blood, hepatic and renal function were reported (Table 2). 25 (18.5%) out of 135 patients showed leukocytosis, an increase in their white blood cell count, as a result of infection. Thrombocytopenia was found in 9 (8.6%) vs. 5 (16.7%) (non-ICU vs. ICU) patients, taking up to 14 (10.4%) in total. Rises in neutrophil level and falls in lymphocyte content were spotted in 64 (47.4%) and 77 (57.0%) of the COVID-19 patients, as a majority of ICU patients showed a sign of neutrophilia (86.7%) or lymphocytopenia (86.7%). All the anomaly in blood cell counts suggested microbial infection and the induced inflammatory response. In parallel, levels of alanine aminotransferase (ALT) or aspartate aminotransferase (AST) were monitored, as 22 (16.3%) or 37 (27.4%) patients showed a surge, reflecting substantial damage to the liver. Mild increase in the concentrations of blood urea nitrogen was observed in 18 (13.3%) patients, suggesting a moderate renal injury. At the same time, an incline in creatinine content was measured for a portion of patients (8.1%), with an insignificant difference between ICU and non-ICU groups. All the blood test results pointed to viral infection as well as possible bacterial co-infection. To present the prognosis between ICU and non-ICU outcomes, we plotted the percentage of each blood parameter in ICU patients vs. that in non-ICU (Figure 2A), where the dotted diagonal line showed a hypothetically equal percentage between two groups. As most points located above the diagonal, those abnormal changes in hematological parameters indicating infection and organ damages, could serve as prognostic biomarkers for COVID-19 patients who might need further intensive care attention.

**Table 2.** Laboratory results of 135 COVID-19 patients upon admission to hospital.

|  | Normal range | Total (n=135) | ICU (n=30) | Non-ICU (n=105) | P value |
| --- | --- | --- | --- | --- | --- |
| White blood cell count, $\times 10^9/L$ | 3.5-9.5 | 6.5 (4.9-8.7) | 10.5 (6.3-14.2) | 6.2 (4.6-7.8) | <0.001 |
| >9.5 |  | 25 (18.5%) | 16 (53.3%) | 9 (8.6%) |  |
| Platelet count, $\times 10^9/L$ | 125-350 | 199.0 (148.0-245.0) | 179.0 (131.8-236.0) | 206.0 (157.5-257.5) | 0.041 |
| <125 |  | 14 (10.4%) | 5 (16.7%) | 9 (8.6%) |  |
| Neutrophil, % | 40-75 | 73.5 (61.1-83.0) | 83.9 (80.7-92.5) | 70.0 (59.8-78.9) | <0.001 |
| >75 |  | 64 (47.4%) | 26 (86.7%) | 38 (36.2%) |  |
| Lymphocyte, % | 20-50 | 17.2 (10.2-27.6) | 7.7 (4.1-11.8) | 20.1 (13.9-30.4) | <0.001 |
| <20 |  | 77 (57.0%) | 26 (86.7%) | 51 (48.6%) |  |
| Alanine aminotransferase, U/L | 9-50 | 24.2 (14.9-39.3) | 33.2 (20.1-72.0) | 22.6 (14.6-32.6) | 0.003 |
| >50 |  | 22 (16.3%) | 11 (36.7%) | 11 (10.5%) |  |
| Aspartate aminotransferase, U/L | 15-40 | 26.7 (20.0-40.7) | 42.6 (26.7-66.0) | 23.4 (18.0-36.9) | <0.001 |
| >40 |  | 37 (27.4%) | 16 (53.3%) | 21 (20.0%) |  |
| Blood urea nitrogen, mmol/L | 2.9-8.2 | 4.7 (3.6-6.3) | 6.1 (4.6-8.4) | 4.5 (3.6-5.4) | 0.001 |
| >8.2 |  | 18 (13.3%) | 8 (26.7%) | 10 (9.5%) |  |
| Creatinine, $\mu\text{mol/L}$ | 40-106 | 65.1 (53.0-82.2) | 67.2 (48.2-86.1) | 64.4 (53.3-81.2) | 0.808 |
| >106 |  | 11 (8.1%) | 3 (10.0%) | 8 (7.6%) |  |
| Troponin T, ng/mL | <0.014 | 0.008 (0.006-0.017) | 0.018 (0.010-0.047) | 0.008 (0.006-0.012) | <0.001 |
| >0.014 |  | 40 (29.6%) | 16 (53.3%) | 24 (22.9%) |  |
| Creatine Kinase Isoenzyme, U/L | 0-25 | 12.6 (10.1-17.0) | 15.5 (11.0-25.4) | 11.9 (9.8-16.1) | 0.006 |
| >25 |  | 15 (11.1%) | 8 (26.7%) | 7 (6.7%) |  |
| Myoglobin, U/L | 25-58 | 29.4 (21.0-63.6) | 63.5 (26.8-166.1) | 26.9 (21.0-47.8) | 0.001 |
| >58 |  | 37 (27.4%) | 16 (53.3%) | 21 (20.0%) |  |
| Lactic dehydrogenase, U/L | 80-285 | 225.0 (182.0-339.0) | 500.5 (303.8-672.5) | 205.0 (177.0-250.5) | <0.001 |
| >285 |  | 43 (31.9%) | 24 (80.0%) | 19 (18.1%) |  |
| Creatine phosphokinase, U/L | 38-174 | 77.0 (53.0-125.0) | 88.5 (50.3-154.8) | 75.0 (53.0-111.0) | 0.381 |
| >174 |  | 20 (14.8%) | 6 (20.0%) | 14 (13.3%) |  |

**Figure 2.**
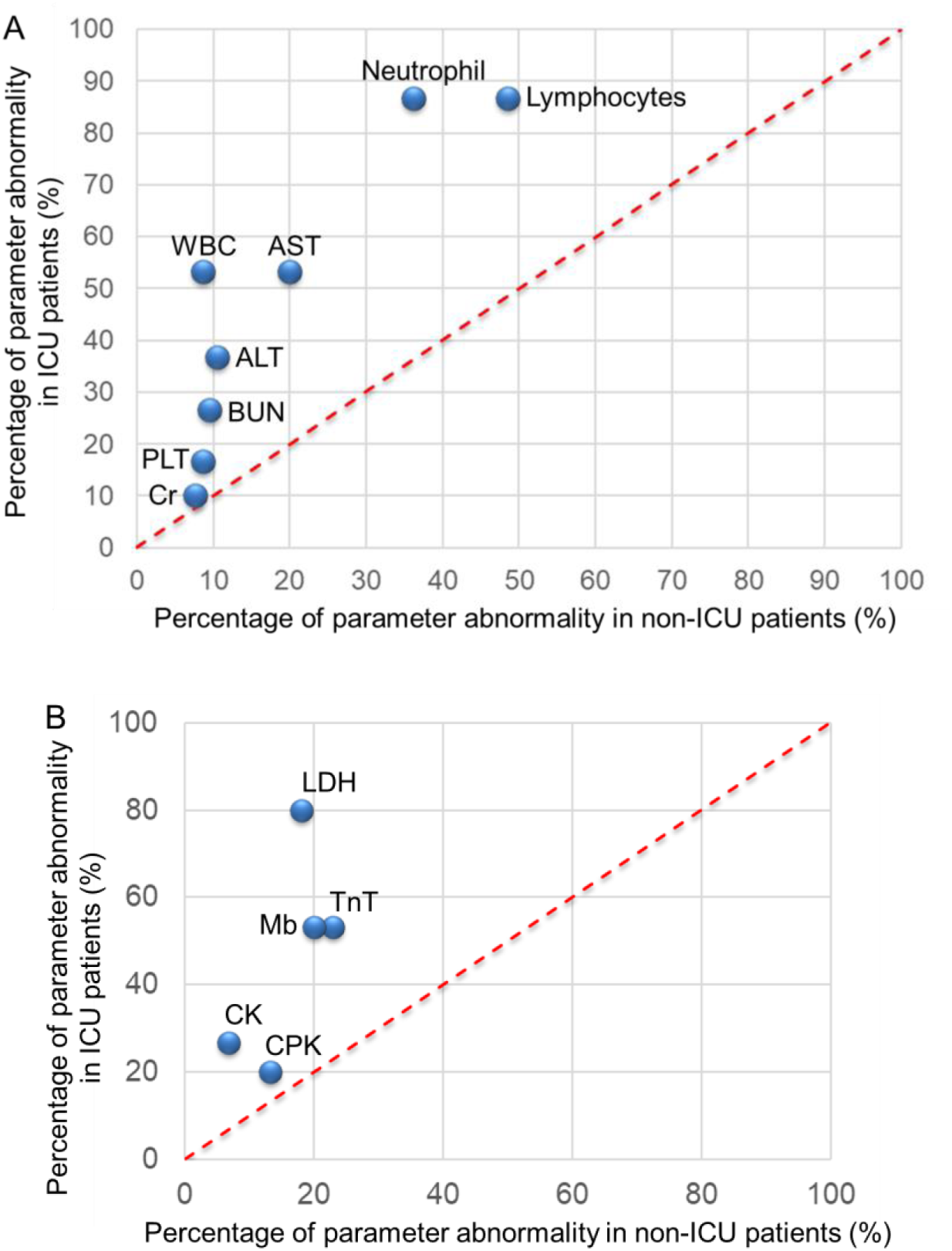
Percentage of each blood parameter as indicated in ICU and non-ICU patients was plotted versus each other. Diagonal (red, dotted) indicated a hypothetically equal percentage between the two groups. (A) Each parameter was labeled as shown, where WBC = White Blood Cells, PLT = Platelet, AST = Aspartate Aminotransferase, ALT = Alanine Aminotransferase, BUN = Blood Urea Nitrogen, Cr = Creatinine. (B) Among blood parameters, data showing myocardial metabolism were shown. TnT = Troponin T, CK = Creatine Kinase Isoenzymes, Mb = Myoglobin, LDH = Lactic Dehydrogenase, CPK = Creatine Phosphokinase.

### Myocardial Parameters Between Mild and Severe COVID-19 Cases

We next examined biochemical markers of myocardial injury for COVID-19 patients, including troponin T (TnT), creatine kinase isoenzymes (CK), myoglobin (Mb), lactate dehydrogenase (LDH), and creatine phosphokinase (CPK) (Table 2). Among 135 patients, 40 (29.6%) patients showed TnT elevation in their sera, including 16 (53.3%) ICU patients and 24 (22.9%) non-ICU patients, as a sign of possible myocardial infraction. 15 (11.1%) patients showed increased levels of CK, where there were 8 (26.7%) cases in the ICU and 7 (6.7%) in the non-ICU, probably suggesting a re-infraction or infract extension. 37 (27.4%) patients had augmented Mb levels with 16 (53.3%) and 21 (20%) patients in the ICU and non-ICU, respectively. Simultaneously, higher LDH and CPK levels were found in 43 (31.9%) and 20 (14.8%) of COVID-19 patients, respectively. Among those patients, LDH content for 24 (80.0%) ICU and 19 (18.1%) non-ICU patients were greater than 285 U/L, whereas CPK level for 6 (20.0%) ICU and 14 (13.3%) non-ICU patients were greater than 174 U/L. All P values for the above parameters (except CPK) demonstrated significantly higher content of those myocardial biomarkers in the ICU patients than in non-ICU, showing a heightened risk of adverse cardiac events in severe COVID-19 patients.

Similarly, in order to predict ICU or non-ICU outcomes, we plotted the percentage of each abnormal blood parameter in ICU patients vs. that in non-ICU patients (Figure 2B), where the dotted diagonal line showed a hypothetically equality between two groups. All points situated within upper diagonal area, suggesting that those abnormal increases in myocardial parameters could be meaningful to assess the prognosis for newly hospitalized COVID-19 patients. Furthermore, the higher above the diagonal line, the higher risk associated with this parameter for a more possibly critical ill case. For instance, patients with high TnT levels predicted a poorer prognosis than those with high CK levels, although both abnormal TnT and CK could serve as prognostic biomarkers for severe COVID-19 disease.

### Radiological Findings for COVID-19 Patients in Different Disease Courses

Following the nucleic acid tests of SARS-CoV-2 pathogens and laboratory tests of blood samples from patients, CT scans were conducted for all patients upon hospitalization. Selected CT images were shown for non-ICU patients, and survival and non-survival groups in ICU patients, where the extent of COVID-19 in different patients could be distinctive (Figure 3). In many a case, bilateral lung involvements were seen at the beginning of infection, although unilateral lung infection was not uncommon. For non-ICU patients, as shown in Figure 3A, at the lower lobe of the right lung located GGOs, together with patchy consolidation and stripy density inside. In Figure 3B, scattered shadows with flocculent density or linear density could be seen in both lungs. For survivors in ICU (Figures 3C-D), diffusive GGO patterns in a typical crescent shape could be observed in both lungs, especially in the subpleural and peripheral areas. At the same time, pulmonary vascular shadows were thickened, suggesting possible obstructions in the patient’ airway. In contrast, for non-survivors (Figures 3E-F), diffusive flocculent density and GGO patterns became overwhelming in the lungs, while curved fluid density shadows were observed in the bilateral thoracic cavity. Flake-like white consolidation were formed as shown in a paving and reticular pattern. Unfortunately, patients with their CT scans shown in Figure 3E-F finally died of respiratory failure on 15 and 24 days, respectively, after those images were taken upon hospital admission.

**Figure 3.**
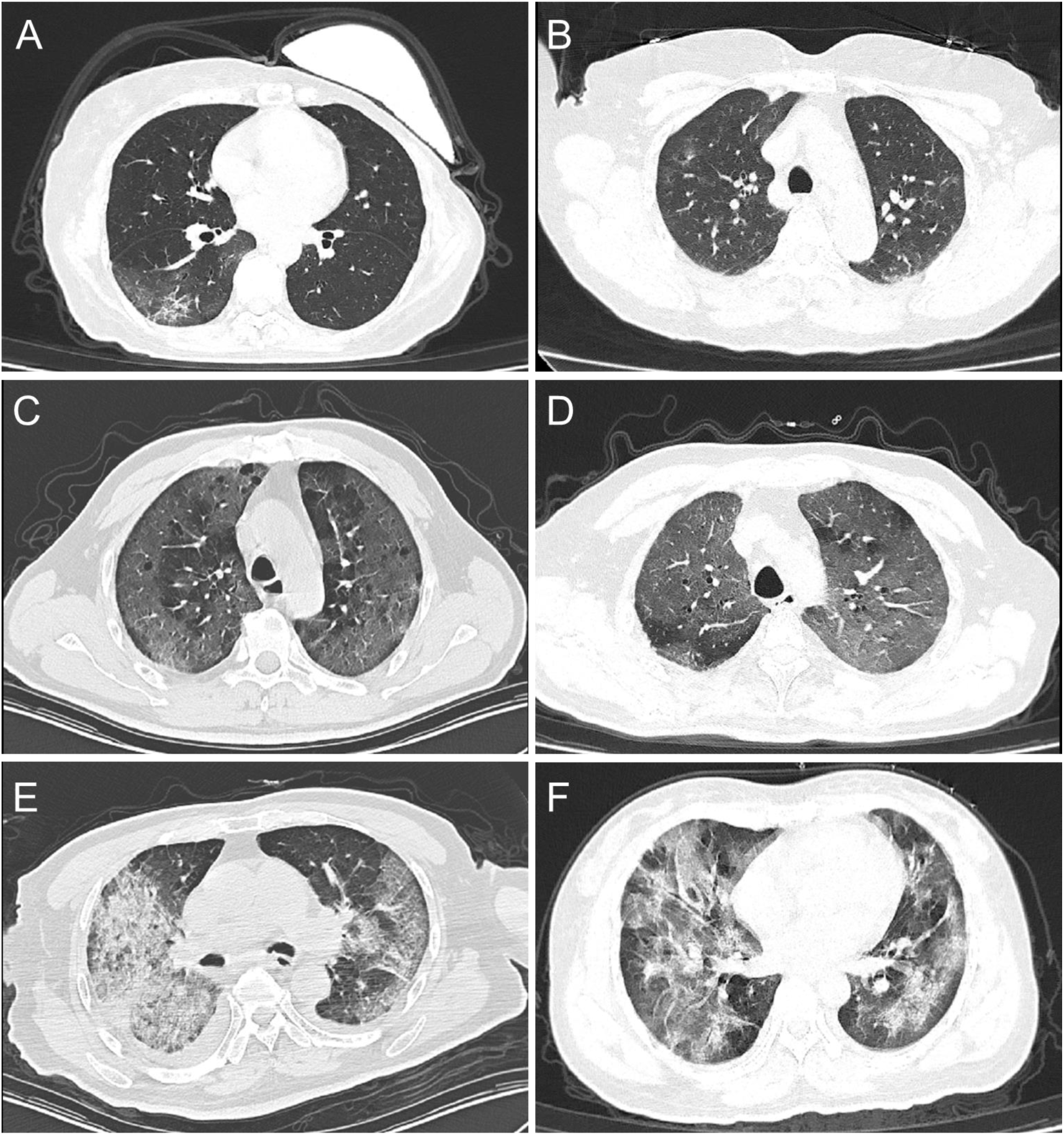
Representative CT images were shown here for non-ICU patients (A-B) and ICU patients (C-F). In ICU patients, (C-D) and (E-F) illustrated CT manifestation for survival group and non-survival group, respectively.

### Myocardial Variables among Severe COVID-19 Patients

As patients in ICU admission were more frequently showing myocardial injury compared to non-ICU patients upon hospital admission, we next investigated whether this occurrence of myocardial anomaly might foretell the eventual outcome of ICU patients in the course of COVID-19 development. For 30 patients in ICU, 18 of them died and 12 survived. All patients had one or more increased myocardial enzymes. Specifically, shown in the following as non-survival vs. survival group (Table 3), 14 (77.8%) vs. 2 (16.7%) in TnT, 8 (44.4%) vs. 0 (0.0%) in CK, 13 (72.2%) vs. 3 (25.0%) in Mb, 14 (77.8%) vs. 10 (83.3%) in LDH, and 5 (27.8%) vs. 1 (8.3%) in CPK. The median values of those myocardial enzyme concentrations (except LDH and CPK) were significantly higher in the non-survival than in the survival (P<0.05). In comparison, there were no significant difference between non-survival and survival groups for all other laboratory parameters (i.e., WBC, AST, etc.). Besides, the factors of patient age, gender and comorbidity did not contribute to make a noticeable difference between survival and non-survival outcome. Relationship between myocardial injury and prognosis for ICU patients was manifested in Figure 4, where percentages of patients with abnormal blood parameters in non-survival and survival group were plotted versus each other. Diagonal line presumed a hypothetically equal percentage in two groups, and around this diagonal were scattered and surrounded by routine parameters including WBC, neutrophils, ALT and AST, etc., which did not show significant difference between non-survival and survival; however, away from the diagonal, all serological parameters representative of abnormal myocardial metabolism (except LDH) in COVID-19 ICU patients portrayed a poor prognosis. The higher above the diagonal, the poorer prognosis to be expected. For example, for COVID-19 patients admitted to ICU, the increased TnT level would suggest worse prognosis than the elevated CPK level, although both parameters could indicate a likely severe diseases development. This was also in line with the findings in Figure 2B, as aberrant TnT values could raise more warning signs than abnormal CPK in the course of disease severity from non-critical to critical conditions.

**Table 3.**
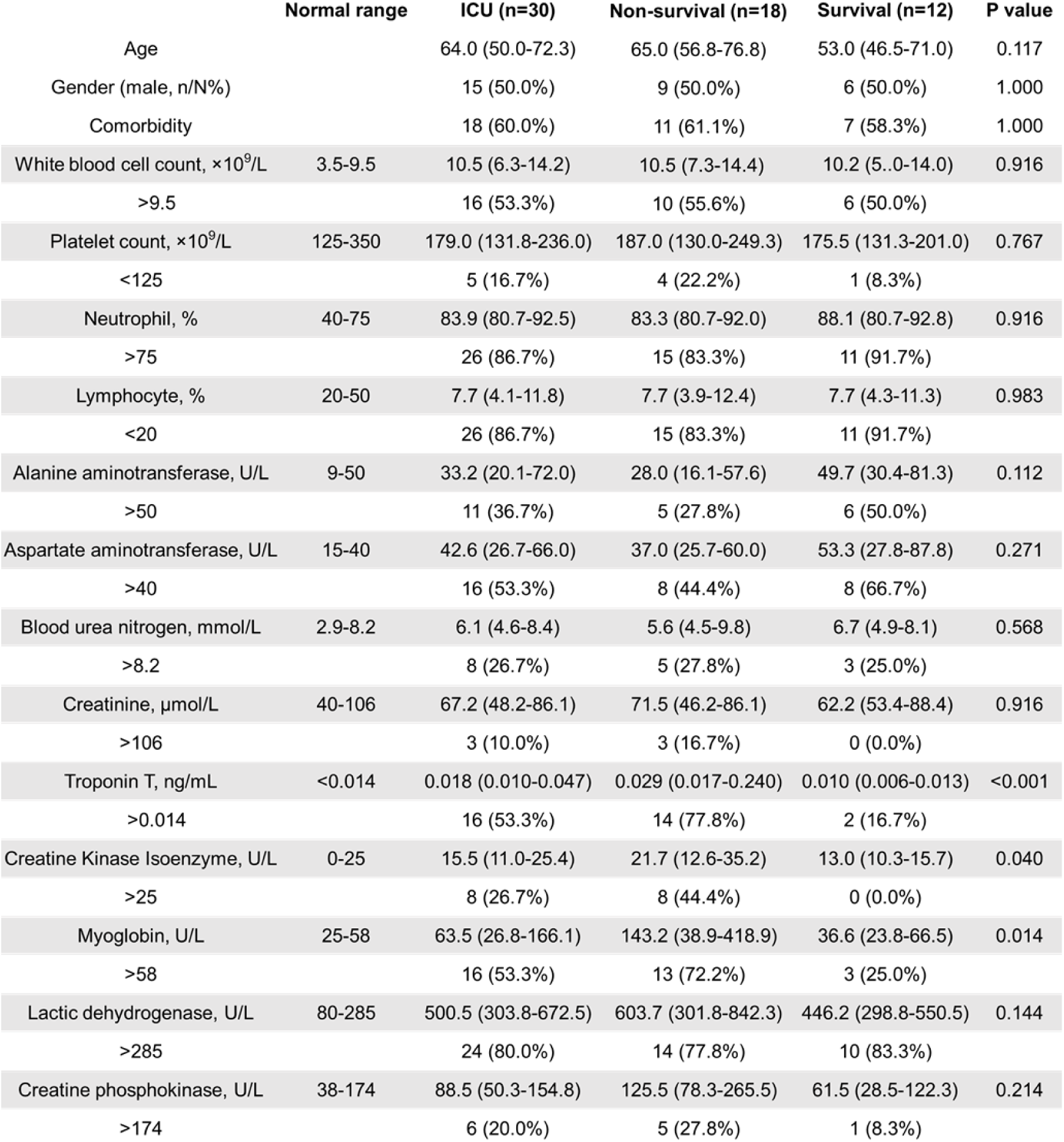
Parameters of COVID-19 ICU patients upon admission to hospital

**Figure 4.**
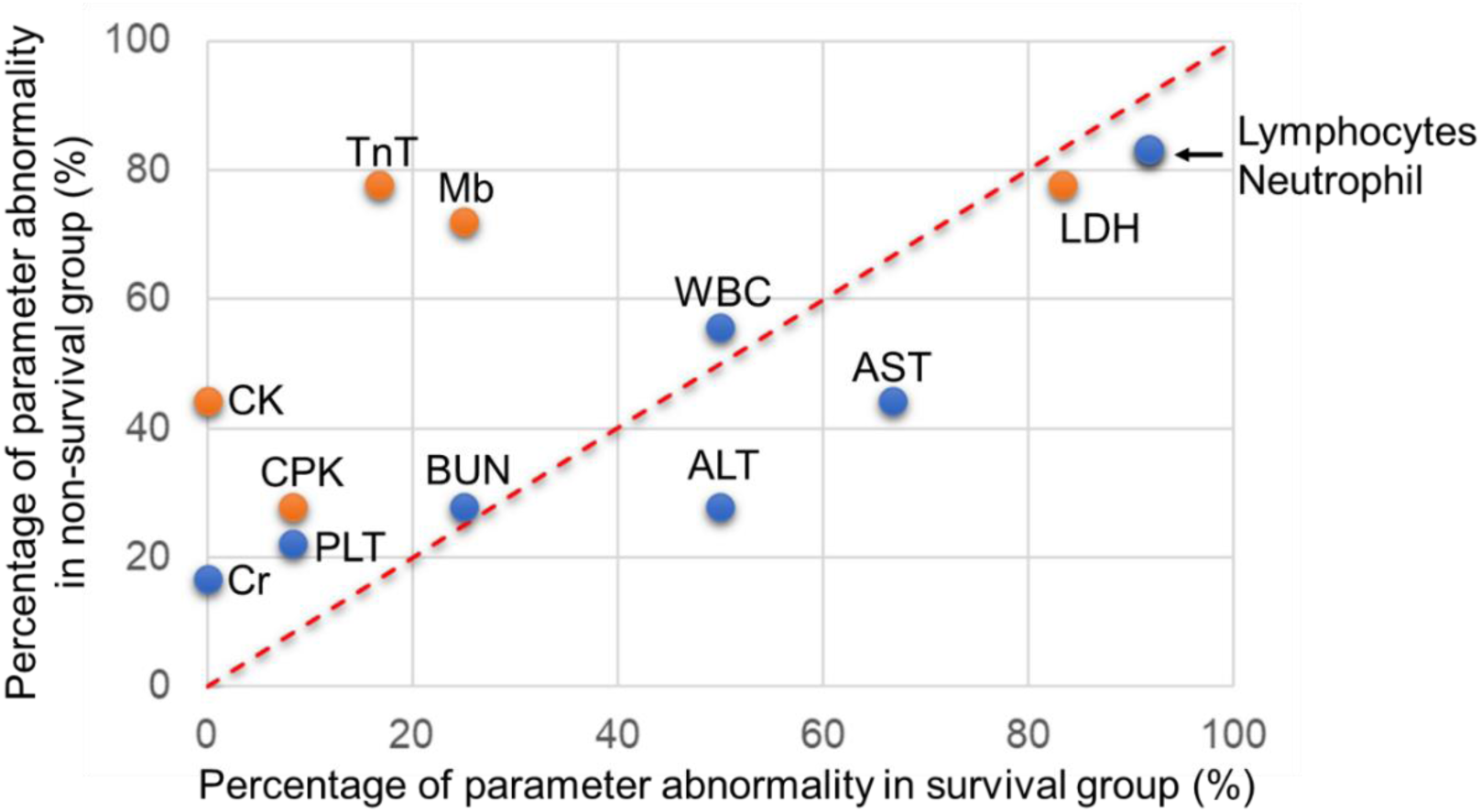
Percentage of each abnormal parameter as indicated in non-survival and survival groups of ICU patients was plotted versus each other. Diagonal (red, dotted) indicated a hypothetically equal percentage between the two groups.

## Discussion

With 95% identity in its *S* gene to SARS-CoV, SARS-CoV-2 oriented its receptor-binding domain (RBD) and optimized its conformation to secure the angiotensin-converting enzyme 2 (ACE2) in the host for cell entry, showing the same manner as SARS-CoV but with higher affinity [9-11]. ACE2 was first known as a vasoconstrictive protein that regulated the renal and cardiovascular function, abundantly expressed in the pneumocytes of lung epithelia and enterocytes of small intestine [12, 13]. This may explain why in addition to the predominant respiratory or pulmonary manifestation, the gastrointestinal symptoms occurred in a substantial portion of COVID-19 patients [14], including diarrhea or vomiting found in our study. This virus entry is further facilitated by a polybasic site containing several arginine residues in *S* protein of SARS-CoV-2, subject to the highly efficient cleavage of a serine protease TMPRSS2 in the target cell, which in turn drives the fusion of viral and host cellular membranes [15, 16].

In previous reports male and elderly group were found susceptible to SARS-CoV-2 [7, 17]. Our study here indicated a minimal difference between different genders of COVID-19 patients in both non-ICU and ICU groups. The fact that male could be more susceptible to COVID-19 may be associated with several non-gender factors, such as smoking history or occupational exposure to the pathogen [18]. Our research confirmed that elderliness posed a high risk of SARS-CoV-2 infection. Many have concluded this to the changing ACE2 activity with aging, where the specific binding of ACE2 to virus outweighs the protective functions of ACE2 to major organs [19].

Our study agreed with others in that hypertension, diabetes and cardiovascular diseases made three leading comorbidities which contributed to the severity of COVID-19 [7, 17, 20]. As ACE2 plays a vital role in regulating blood pressure and renal function, chronic disease development and long-term medication may confound the ACE2 activity in each individual [21]. Meanwhile, SARS-CoV-2 infection may lead to the downregulation of ACE2 expression in a similar manner as SARS-CoV did, therefore exacerbating organ damages [22]. However, the association of COVID-19 transmissibility with ACE2 activity remains uncertain, while high frequency of those chronic diseases among population, interlinked pathogenesis between those diseases, and compromised immune systems in those patients of comorbidity, could make them vulnerable to the viral infection.

COVID-19 patients were found to have higher viral load in the nasal swabs or sputum samples than in the throat swabs [23, 24]. Moreover, SARS-CoV-2 was inclined to infect the lower airway, including trachea, bronchi, and alveoli [17]. This agreed with our findings where involuntary cough was the most common symptom in COVID-19 patients, followed by febrile illness as a sign of infection. All those results pointed out that in the quest to prevent COVID-19 contraction and stop SARS-CoV-2 spreading, oronasal covering or social distancing is an imperative measure to thwart the otherwise respiratory tract transmission intra- or inter-personally, or it would become harder to contain the virus for avoiding further deep lung infection.

Once inhalation of SARS-CoV-2 contained particles delivers virus to the lung, where alveolar epithelia actively expressed ACE2 proteins, it further induces harmful lung injuries [12]. In our study, CT images for COVID-19 patients of different disease developments pointed out pathological changes in infected lungs, from intensification of patchy GGO, to compressed consolidation, and to formed pave pattern. Thereby the gas exchange could be gradually impaired at the interface of alveolo-capillary membrane, leading to fluid filling in alveolar units and eventual pulmonary edema, which posts a proof of severe pneumonia and a warning of respiratory failure. This echoes other radiological findings in COVID-19 patients where 61.1%-85.0% critically ill patients did not survive ARDS after mechanical ventilation [7, 20, 25-27]. In our study 15 (83.3%) out of 18 COVID-19 deceased cases suffered and died from respiratory failure (data not shown). Postmortem biopsy of one COVID-19 patient showed evident ARDS signs in both lungs, represented by diffuse alveolar damages, hyaline membrane formations, and viral cytopathic changes in the intra-alveolar space [28]. This pathological finding correlates the prevalence of ARDS-caused mortality in COVID-19 patients and postulated a possibility of viral invasion in deep lungs.

Oronasal entry of SARS-CoV-2 led to its direct infection of pulmonary and digestive systems, as well as earning a chance to enter bloodstream and then contract extrapulmonary organs through blood flow. Upon hospital admission, our clinical data from a substantial portion of COVID-19 patients showed the abnormality in several blood parameters, including peripheral blood cells, hepatic enzymes, renal metabolites and myocardial proteins, indicating acute assaults to immune systems together with damages to major organs including liver, kidney and heart. All parameters (except Cr and CPK) showed significant difference between ICU and non-ICU patients, establishing them as prognostic markers for status of COVID-19 severity. This result is in concert with previous reports [1, 20]. However, none of those parameters suggested a difference between survivors and non-survivors in ICU group, except myocardial markers including TnT, CK and Mb. This finding here recommended that upon hospitalization the myocardial enzyme levels could serve as prognostic indicators for in-hospital COVID-19 severity and mortality even before medical care is given.

Regardless of previously underlying diseases, a considerable amount of cardiac injury in COVID-19 patients have been noticed with correlation to disease severity and mortality [29]. In our view, two pathways could be both involved, each to a different extent. Firstly, a direct viral invasion by riding the blood flow is plausible, a process mediated by ACE2 that is highly and specifically expressed in the pericytes of heart [30]. Study has revealed that SARS-CoV genome was detected in 35% autopsied heart samples of 20 SARS patients, concurrent with lowered ACE2 activity, inundated macrophages, and significantly shortened illness durations before death [31]. This learning offers a tip for understanding SARS-CoV-2 infection. Secondly, SARS-CoV-2-triggered hypercytokinemia (more well-known as cytokine storm) may result in multiorgan damages, making an indirect but deadly hit to heart [29]. As a proof, activations of both T-helper-1 (Th1) and T-helper-2 (Th2) cells were identified in COVID-19 patients, and elevated cytokine levels were found in ICU patients compared to non-ICU ones, such as interleukin-2, -7, and tumor necrosis factor α [1]. While a cascade of cytokine release was initiated, it also led to dysregulation of immune systems, promoting cardiomyocyte death and eventually heart failure. Insofar, biopsied heart sample from one COVID-19 patient did not endorse a direct cardiac impairment of SARS-CoV-2, where overactivation of Th17 and cytotoxic T cells was found in the peripheral blood, voting for an indirect heart injury [28]. More evidences are necessitated to explore the underlying mechanism of organ impairment by SARS-CoV-2, which could inspire effective treatments to decelerate and stop the increasing COVID-19 fatality.

In summary, we here highlighted myocardial variables from 135 COVID-19 patients prior to their in-hospital treatment, and utilized them as prognostic indicators for disease severity and mortality of COVID-19, with a hope to assist decision-making of clinicians when treating patients.

## Data Availability

Data access will be provided upon request once approved.

## Notes

### Author contributions

JiaZ, DD and ZT conceived the idea and designed the study. JiaZ, DD, CC, and ZT had approved access to all data in the study. JiaZ, CC, JinZ, XH, GL, WX and ZT contributed to writing of the report. PF and ZT contributed to the figure preparation. JiaZ, XH and ZT contributed to the statistical analysis. JiaZ and ZT took responsibility for the integrity of data and the accuracy of the data analysis. All authors contributed to data acquisition and analysis, and all reviewed and approved the manuscript submission.

